# A COVID-19 Model for Local Authorities of the United Kingdom

**DOI:** 10.1101/2020.11.24.20236661

**Authors:** Swapnil Mishra, Jamie Scott, Harrison Zhu, Neil M. Ferguson, Samir Bhatt, Seth Flaxman, Axel Gandy

**Author notes:** Contributed equally (Swapnil Mishra, Jamie Scott, Axel Gandy).

## Abstract

We propose a new framework to model the COVID-19 epidemic of the United Kingdom at the level of local authorities. The model fits within a general framework for semi-mechanistic Bayesian models of the epidemic, with some important innovations: we model the proportion of infections that result in reported deaths and cases as random variables. This is in contrast to standard frameworks that model the latent infection as a deterministic function of time varying reproduction number, *R*_*t*_. The model is tailored and designed to be updated daily based on publicly available data. We envisage the model to be useful for now-casting and short-term projections of the epidemic as well as estimating historical trends. The model fits are available on a public website, https://imperialcollegelondon.github.io/covid19local. The model is currently being used by the Scottish government in their decisions on interventions within Scotland [1, issue 24 to now].

## 1 Introduction

The United Kingdom (UK), with 53,775 COVID-19-attributes deaths as of 20^*th*^ November 2020 is fifth in the global tally, after the United States, Brazil, India and Mexico [2]. Initially, after implementing the first nationwide lockdown on 23^*rd*^ March 2020, UK made significant progress in containing the SARS-CoV-2 epidemic, pushing the time-varying reproduction number *R*_*t*_ below 1 [3]. However, with a recent surge in epidemics [4], UK went into a second national lockdown on 5^*th*^ November 2020 [5]. This second wave has seen both an increase in prevalence and rate of growth of infections across UK [4]. The health care system burden has increased considerably to a 7 day rolling average of almost 1750 new admissions daily, as of 20^*th*^ November 2020 [6].

However, the UK has seen localised outbreaks in the time period between the initial announcement of easing of lockdown on 11^*th*^ May [7] and the announcement of the second lockdown. For instance, Leicester [8] saw an increase in infections after the initial announcement of easing of lockdown on 11^*th*^ May, just before the period when rest of the country was about to see further relaxations on 4^*th*^July 2020 [9]. Similar increases in other parts of England occurred and a tier system [10] was introduced on 12^*th*^ October 2020. As most of the surveillance system currently in place for the UK produce either nation or region-specific estimates [11], this tier system was mainly a retrospective action rather than a preventive action.

This underscores the need for early identification of outbreaks at local and regional levels. We hope local outbreaks can be identified and managed by continuously monitoring the available health care data from various public health agencies across the UK. Early identification of outbreak and knowledge of the direction of the epidemic at a local level can help target measures such as enhancing local testing capacity, closing and quarantining specific areas, enforcing workplace safety measures, and increasing community outreach campaigns to promote adherence to social distancing and increasing participation in Test and Trace. Effective local measures could help avoid national lockdowns while minimising the spread of the epidemic, essential for reviving health care services, economy and society at large.

To identify trends and facilitate the identification of outbreaks at the local level, we implement a semi-mechanistic Bayesian transmission model for SARS-CoV-2 at local authority levels for the UK. The model aims to assess and project the evolution of the epidemic at a local level and to provide estimates for the time-varying reproduction number for local areas of the UK.

While identifying a local epidemic, two factors need to be accounted for: the current number of individuals that are infected/infectious and the rate of transmission of the virus, captured in the time-varying reproduction number *R*_*t*_. We use the projected cases in 2 or 3 weeks from the current point in time as a measure to identify the possible areas for concern. Our proposed approach combines the two key factors into a single meaningful measure. Our projection does not take any interventions and changes in testing strategies into account that may be enacted in the future, hence giving an estimate of the evolution of the local epidemic in the current situation.

We have extended the Flaxman *et al*. [12] model for each local authority by incorporating the following innovations:

- The model incorporates the reported cases for each local authority from June 1^*st*^ 2020.
- Survey data from the Office of National Statistics [13] and from the Real-time Assessment of Community Transmission study [14] is used to calibrate our estimates of true infections.
- The model incorporates a time-varying infection fatality rate (fraction of infections that leads to deaths) and infection ascertainment rate (fraction of infections identified as positive cases).
- The infection process is random, i.e., it is no more a deterministic function of time varying reproduction number and previous infections given a serial-interval (generation distribution). This is to better account for variability in areas that have low infection numbers.

Regularly updated results from the model are presented at covid19local.^1^ The model is currently being used by the Scottish government in their response to the epidemic [1, issue 24 to now].

## 2 Methods

### 2.1 Data

We combine data from national statistics and public health bodies across the UK. Table 1 lists all the sources we use. For reported cases, the model uses the date on which the specimen was collected. To account for reporting variations within a week, we aggregate case and deaths data to be weekly. We do not use the last three days of data while fitting our model, to account for reporting delays.

**Table 1:**
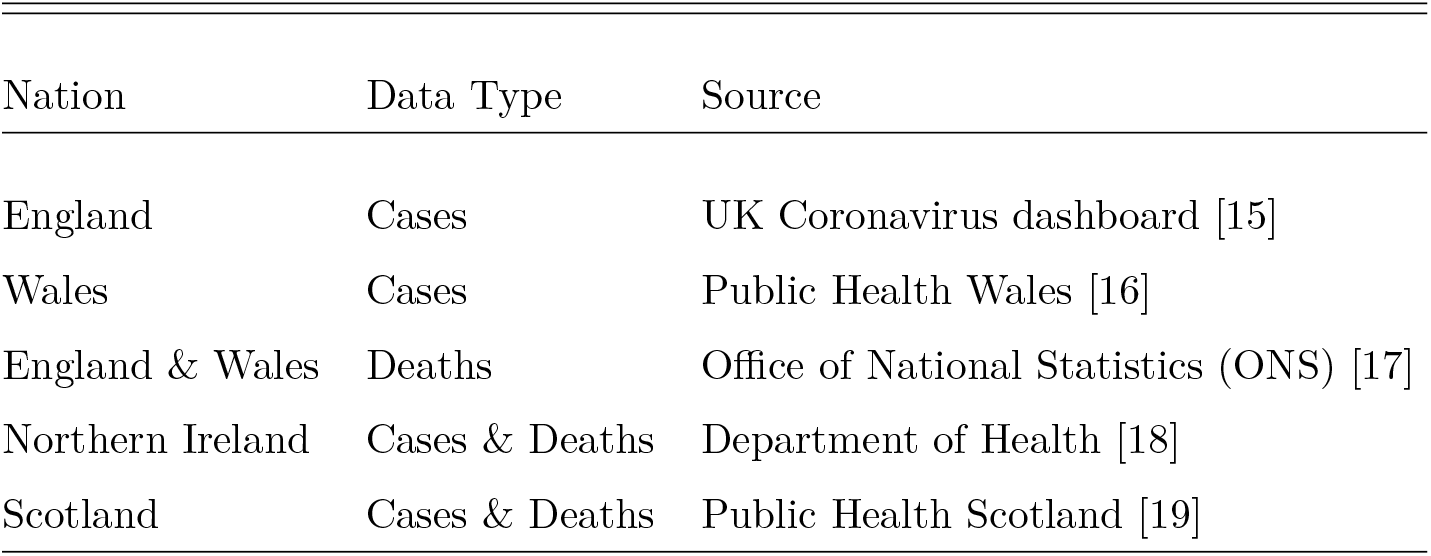
Sources for various types of data on COVID-19 for nations within the UK.

| Nation | Data Type | Source |
| --- | --- | --- |
| England | Cases | UK Coronavirus dashboard [15] |
| Wales | Cases | Public Health Wales [16] |
| England & Wales | Deaths | Office of National Statistics (ONS) [17] |
| Northern Ireland | Cases & Deaths | Department of Health [18] |
| Scotland | Cases & Deaths | Public Health Scotland [19] |

Furthermore, we use the weekly COVID-19 infection survey conducted by the ONS [13], and the findings from the Real-time Assessment of Community Transmission (REACT) study [14]. Data from these nation-wide surveys are used for calibrating the estimates of infections produced by our model.

### 2.2 Model

Flaxman *et al*. [12] introduced a Bayesian semi-mechanistic model for estimating the transmission intensity of SARS-CoV-2. The model is based on the renewal equation [20], using the time-varying reproduction number *R*_*t*_ to generate new infections. In the early part of the SARS-CoV-2 epidemic in the UK, reported case data is unreliable, so the original formulation of the model relied on observed deaths data and calculated backwards to infer the true number of infections. The latent daily infections are modelled as the product of *R*_*t*_ with a discrete convolution of the previous infections, weighted using an infection-to-transmission distribution specific to SARS-CoV-2 [21].

We modify the original Bayesian semi-mechanistic model and apply it to all local authorities (LA) of the UK to infer the reproduction number over time (*R*_*t*_) and the daily infections. We have extended the original model in the following ways:

Interventions are not being explicitly included in the local models. Instead, a weekly random walk on *R*_*t*_ is used.

We include the reported number of cases for each LA from June 1^*st*^ 2020 and observed deaths during the entire epidemic. We parametrise *R*_*t*_ with a random effect for each week throughout the epidemic for each LA separately (no joint inference of parameters across different local authorities). The weekly random effects are encoded as a random walk with normally distributed updates, where at each successive step the random effect has an equal chance of moving upwards or downwards from its current value. Our model is implemented using epidemia^2^ [22], a general purpose R package for semi-mechanistic Bayesian modelling of epidemics.

Fitting reliable and robust models for local areas is a challenging task as raw counts for local areas can be low. To reliably estimate model parameters for all local authorities, we take a three-stage top-down approach for fitting our LA models.

To calibrate parameters in our models, specifically the infection fatality ratio (ifr) and the infection ascertainment rate (iar), we first fit a model to the whole of England using weekly deaths and case data, as well as survey data from ONS and REACT. For all subsequent individual models, we use these estimated ifr and iar values as the prior. The estimated ifr and iar values are shown in Figure 1.

**Figure 1:**
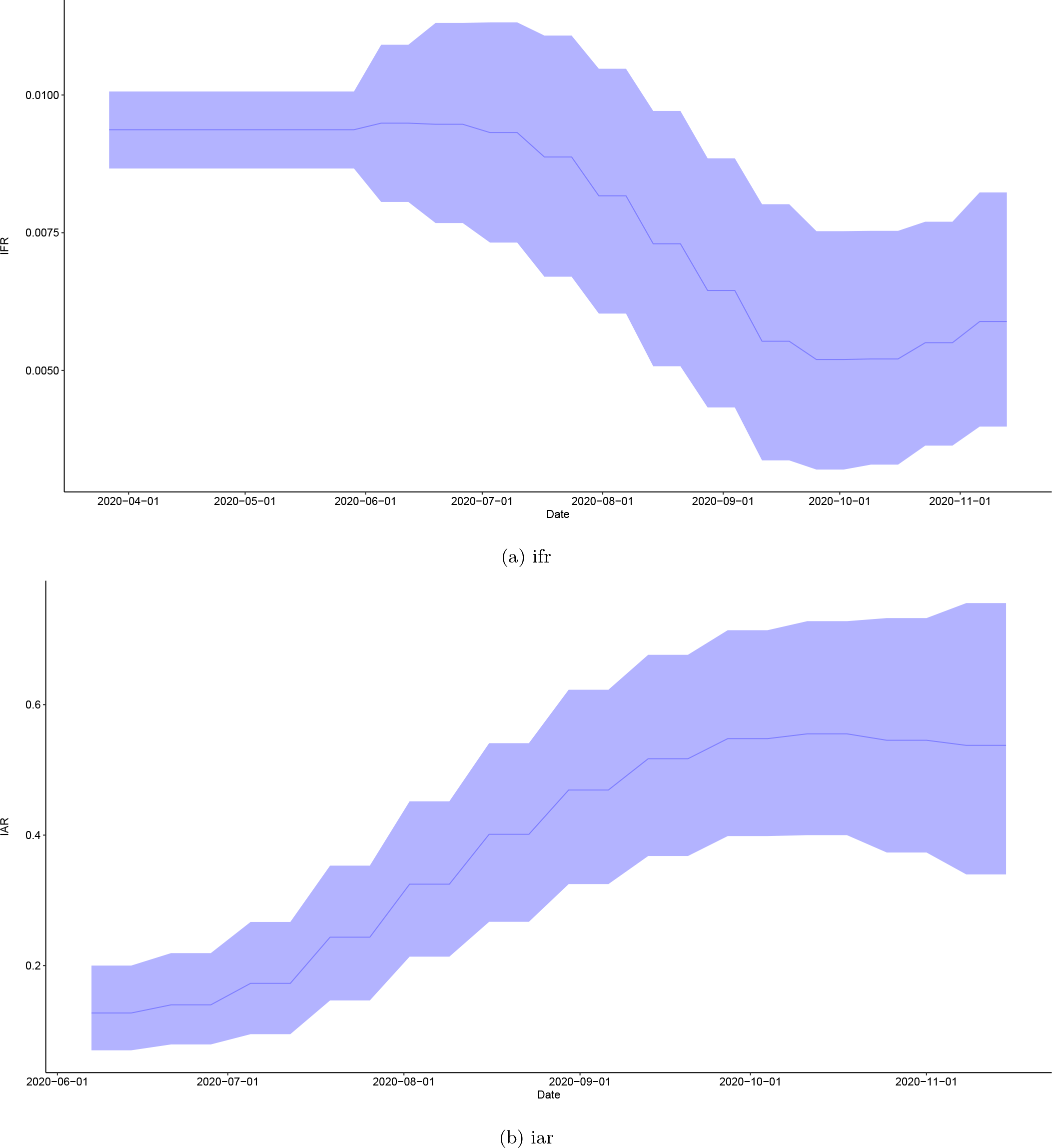
Time varying estimates of the infection fatality ratio (ifr), figure (a), and the infection ascertainment rate (iar), figure (b), for England. The solid line is the mean estimate and the filled area denotes 90% pointwise credible intervals.

In the next step, we fit individual models to Northern Ireland, Scotland and Wales and to the 9 regions of England by using aggregated deaths and cases data, along with the estimated prior values for ifr and iar. The estimates from these 12 regions provide us with underlying trends of *R*_*t*_ for local areas within a particular region. We report the results of these regions, together with estimates for England and the UK as a whole.

Finally, we fit individual models for each of the local areas. The *R*_*t*_ for a local area is parametrised using a weekly random walk and the estimated *R*_*t*_ of the region the local area is situated in. Using a background *R*_*t*_ trend helps us to stabilise our inference procedure for local areas. The background *R*_*t*_ of the regions is not used for last 45 days to make sure trends in a local area are driven mainly by the data from the local area.

For implementation details please look at the infections_normal^3^ branch of epidemia [22].

## 3 Results

### Time varying reproduction number (*R*_*t*_)

Figure 2 shows the geographical variation in the posterior probability that *R*_*t*_ is greater than or less than 1 for all the local areas in the UK. There is substantial geographical clustering; most areas in Wales, Northern Ireland, and the North West are showing either a decrease or likely decreasing tendency in new infections. We summarise the results for various nations in the UK and regions of England in Table 2. We include estimates of *R*_*t*_ and infections over time for each local area on our daily updated website^4^ [23].

**Table 2:** Table with estimated R values and its probability of being greater than 1 for all nations of UK and regions of England as of 2020-11-14.

| Area | Date | P($R > 1$) | $R$ |
| --- | --- | --- | --- |
| Northern Ireland | 2020-11-14 | 0.17 | 0.79[0.5-1.21] |
| North West | 2020-11-14 | 0.19 | 0.8[0.49-1.26] |
| Wales | 2020-11-14 | 0.21 | 0.79[0.45-1.23] |
| Scotland | 2020-11-14 | 0.24 | 0.82[0.49-1.29] |
| Yorkshire and Humber | 2020-11-14 | 0.43 | 0.96[0.6-1.42] |
| UK | 2020-11-14 | 0.47 | 0.98[0.64-1.39] |
| England | 2020-11-14 | 0.48 | 0.99[0.63-1.45] |
| North East | 2020-11-14 | 0.49 | 0.99[0.65-1.44] |
| East Midlands | 2020-11-14 | 0.56 | 1.04[0.68-1.54] |
| West Midlands | 2020-11-14 | 0.70 | 1.15[0.73-1.66] |
| South West | 2020-11-14 | 0.71 | 1.14[0.75-1.64] |
| London | 2020-11-14 | 0.73 | 1.14[0.77-1.63] |
| South East | 2020-11-14 | 0.84 | 1.24[0.84-1.75] |

**Figure 2:**
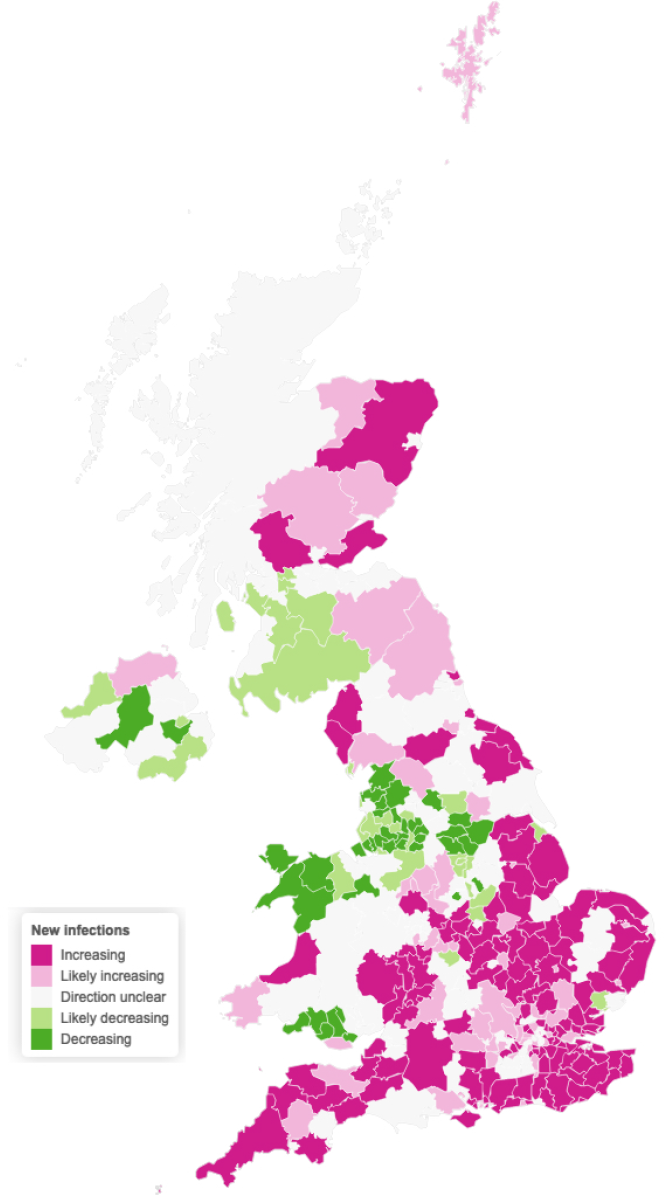
Estimates of the probability that the time-varying reproduction number *R*_*t*_ is greater than or less than one in each local area.. We consider an area to have increasing new infections if our model estimates that the reproduction number R is greater than 1 with probability of at least 90%. Likely increasing indicates a probability between 75% and 90%. Decreasing and likely decreasing are defined analogously, but consider R less than 1. These estimates are as of 2020-11-14.

### Hotspots

We have used our model to project the course of the epidemic in future. We argue that to develop a surveillance system for local areas just looking at current case counts or current *R*_*t*_ is not enough. Looking at only one value can give a misleading impression about the evolution of the epidemic. A low case count with high *R*_*t*_ is dangerous as exponential growth can lead to a sudden increase in infections/cases, and similarly as low *R*_*t*_ combined with a high number of infections can also lead to a substantial epidemic. Hence, we combine both current infections and *R*_*t*_ to predict the cases in future 2/3 week period. This way we account for the interaction between infections and *R*_*t*_ to a single metric that is easily understood or can be acted upon.

Furthermore, instead of just predicting case counts which at times can lead to very high or low numbers, we summarize this count by probabilities, which essentially gives us an estimate of how certain we are that a given area will exceed a given set threshold in the projection period. Specifically, an area is defined as being a hotspot if the two/three week prediction of cases per 100K population are projected to exceed a given threshold, e.g. 200 cases. Figure 3 shows the hotspot probability for all the local areas in the UK for the period 2020-11-29 to 2020-12-05.

**Figure 3:**
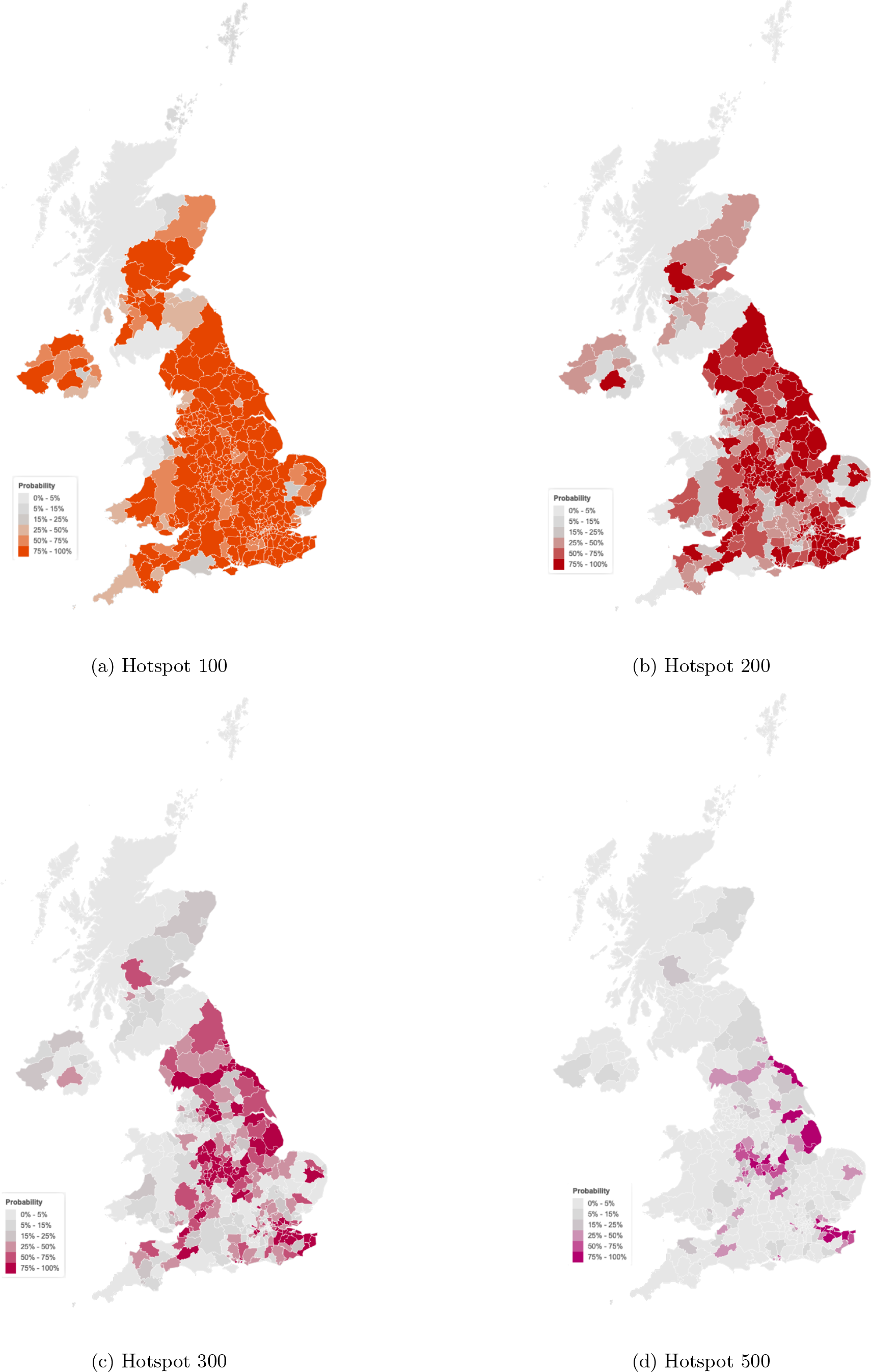
Probability of local authority areas having more than 100, 200, 300 or 500 cases per 100K (2020-11-29 to 2020-12-05)

## 4 Limitations

Major model assumptions of our model include:

- Assuming a fixed probability distribution with fixed mean and coefficient of variation for the delays from infection to onset of symptoms and from symptom onset to death
- Assuming a fixed probability distribution with fixed mean and coefficient of variation for the generation time distribution
- Assuming the generation time and serial interval distributions are the same
- Assuming average dynamics locally and across all ages
- Projections made by our model assumes no change in current interventions (lockdowns, school closures, and others) in the local area beyond those already taken about a week before the end of observations.
- Increases in testing can account for increases in cases; currently the model does not account for this.
- Each local area is treated independently of each other. Thus the epidemic in a region is neither affected by nor affects any other region. It also does not include importations from other countries.
- Changes in measures and lockdowns are not an explicit part of the model; hence their effect may not appear for 1-2 weeks post the date of implementation.

Further limitations of the model setup are discussed in [12].

## Data Availability

The data in this study is all public data. For details got to the url

https://imperialcollegelondon.github.io/covid19local/#details

## Footnotes

1 https://imperialcollegelondon.github.io/covid19local

2 https://imperialcollegelondon.github.io/epidemia

3 https://github.com/ImperialCollegeLondon/epidemia/tree/infections_normal

4 https://imperialcollegelondon.github.io/covid19local/

